# Assessment of the Performance of Expired and Unexpired BinaxNOW COVID Rapid Antigen Test Kits Using Positive and Negative Controls

**DOI:** 10.1101/2023.03.01.23286637

**Authors:** Mary Jane E. Vaeth, Asia Mitchell, Kristie Sun, Maisha Foyez, Lourdes Byrd, Maryam Elhabashy, Minahil Cheema, Ishaan Gupta, Sophia Purekal, Charles Locke, Melinda Kantsiper, Zishan Siddiqui

## Abstract

Rapid antigen tests are widely used to diagnose infection with SARS-CoV-2, and millions of kits have been distributed for free by government agencies. However, unused and expired kits beyond their final expiration dates remain prevalent in people’s homes. This study aimed to determine the accuracy of expired BinaxNOW COVID rapid antigen test kits. 100 expired and 100 unexpired test kits were checked for sensitivity and specificity using positive and negative controls, respectively. The results showed that there was no change in the sensitivity and specificity of BinaxNOW COVID rapid antigen test kits four months beyond the manufacturer-extended expiration date when using manufacturer-provided positive controls. The findings provide confidence in the accuracy of expired test kits, which could potentially reduce waste and strengthen supply chain resilience for pandemic preparedness. Further research utilizing actual human specimens can help determine the true accuracy of expired rapid antigen test kits in clinical use.

## Background

Rapid antigen tests have been widely used to diagnose infection with SARS-CoV-2. Tests are now easily accessible, thanks in part to the hundreds of millions of kits distributed for free by government agencies (Abbott, 2021). Despite stability testing determined expiration date extensions of up to 16 months for some of the kits by the manufacturer, unused and expired kits beyond their final expiration dates remain prevalent in people’s homes. (Deane, The New York Times, 2021). People may still be using expired kits beyond any extension due to convenience or ignorance of the true expiration date of a kit in their home. Clinicians need guidance on interpreting and determining the appropriate course of action if a patient tests positive using an expired COVID-19 rapid antigen test. Extending the expiration date to its fullest usable limit would reduce waste, control costs, and strengthen supply chain resilience for pandemic preparedness. In light of this, it is crucial to assess the accuracy of expired test kits beyond manufacturer extensions, as some may still be usable.

## Methods

We sought to determine the accuracy of BinaxNOW COVID rapid antigen test for expired test kits. 100 expired and 100 unexpired test kits were checked for sensitivity using positive controls from the manufacturer, and the differences were calculated. The expired test kits used in the study had been expired for 4 months (including additional manufacturer extensions) at the time of the study. Positive controls are swabs with samples known to contain a specified quantity of the target analyte, in this case, the SARS-CoV-2 virus nucleocapsid protein. Positive controls serve to verify the performance characteristics of a diagnostic test and ensure proper functioning of the test itself. Further technical details about positive control are unavailable, cited as proprietary by the manufacturer. 50 expired and unexpired tests were evaluated and compared for specificity using unused sterile swabs as negative control. Sample processing followed the manufacturer’s instructions.

**Table:** Agreement Matrix for Expired and Unexpired Rapid Antigen Test with Positive and Negative Controls

|  | Unexpired Kit |  |  |  |
| --- | --- | --- | --- | --- |
|  | Using Positive Control |  | Using Negative Control |  |
|  | Positive | Negative | Positive | Negative |
| Expired Kit |  |  |  |  |
| Positive | 100 | 0 | 0 | 0 |
| Negative | 0 | 0 | 0 | 50 |

## Results

The results of the study showed that 100% of both expired and unexpired test kits tested positive when the positive control was used yielding a sensitivity of 100% for each group showing a difference of 0% (95% CI -3.7%. to 3.7%) Additionally, all negative controls tested negative, further demonstrating a specificity of 100% for both the expired and unexpired test kits (Difference of 0% [95% CI -7.1% to 7.1%]).

## Discussion

This study using manufacturer-provided positive controls found no change in BinaxNOW COVID rapid antigen test sensitivity and specificity 4 months beyond the manufacturer-extended expiration date, confidence in expired kit accuracy. However, this study was limited by the use of positive controls without real-world specimens, which may contain broader variability in viral load. Further research utilizing actual human specimens can help determine the true accuracy of expired rapid antigen test kits in clinical use. Reliable testing remains vital in the COVID-19 response as health systems transition to long-term strategies and sustainable supply chains. Understanding the practical shelf life of expired kits will help with clinical decision making, responsible resource management, reduced waste, and maintaining readiness for future outbreaks.

## Data Availability

All data produced in the present work are contained in the manuscript.

## References

Abbott. (2021, January 12). Abbott announces fulfillment of federal government purchase of 150 Million BinaxNOW™ covid-19 rapid tests and is now ready to support commercial distribution. Abbott MediaRoom. Retrieved February 4, 2023, from https://abbott.mediaroom.com/2021-01-12-Abbott-Announces-Fulfillment-of-Federal-Government-Purchase-of-150-Million-BinaxNOW-TM-COVID-19-Rapid-Tests-and-Is-Now-Ready-to-Support-Commercial-Distribution

Deane, E. (2022, January). BinaxNOW COVID-19 AG Card product expiry update. Retrieved February 4, 2023, from https://www.healthvermont.gov/sites/default/files/documents/pdf/COVID-19-BinaxNOW-Expiry-Extension.pdf

The New York Times. (2021, January 20). How to Read a Covid Test Result: The Basics. Retrieved from https://www.nytimes.com/2021/01/20/health/covid-test-result.html

World Health Organization. (2022, February 1). Tonnes of COVID-19 health care waste expose urgent need to improve waste management systems. Retrieved from https://www.who.int/news/item/01-02-2022-tonnes-of-covid-19-health-care-waste-expose-urgent-need-to-improve-waste-management-systems

